# Household transmission in people infected with SARS-CoV-2 (COVID-19) in Lima-Perú

**DOI:** 10.1101/2020.09.06.20189456

**Authors:** Yolanda Angulo-Bazán, Gilmer Solis-Sánchez, Joshi Acosta, Fany Cardenas, Ana Jorge, Cesar Cabezas

**Author notes:** Master in Public Health (Joshi Acosta). Doctor of Medical Microbiology (Joshi Acosta).

## Abstract

**Objective:** Describe the characteristics of SARS-CoV-2 infection among household members with a confirmed primary case of COVID-19 in low burden districts in Metropolitan Lima.

**Materials and Methods:** A retrospective, secondary database review study was conducted. The information was collected from an epidemiological surveillance activity in close contacts (co-inhabitants) in 52 households in Metropolitan Lima with only one member with COVID-19. A reevaluation was carried out in 10 households. Epidemiological and clinical variables were evaluated and its association with the result of the rapid serological test (presence of IgG, IgM or both).

**Results:** Secondary cases were found in 40 households, which represents an average of 49.9% identification per household. A secondary attack rate of 53.0% (125 cases) was found among cohabitants, with 77.6% of cases being symptomatic (symptomatic / asymptomatic ratio: 3.5). The presence of fever and / or chills was found in 40.0% of people with a positive result, followed by a sore throat, in 39.2%. Ageusia and anosmia were present in 22.4% and 20.8% of cases, respectively. A reevaluation in 40 family members 33.6 ± 2.7 days after the first evaluation, show the persistence of positive IgM and IgG in the 20 positive cases in the first evaluation.

**Conclusion:** Having a primary case of COVID-19 in home, the secondary attack rate of this infection is 53%; however, in a significant proportion of households evaluated there was no positive case, beyond the primary case. The epidemiological and clinical characteristics found in this case were in accordance with what has already been reported in other international series.

## INTRODUCTION

The SARS-CoV-2, is a RNA virus that belongs to the family of the *Orthocoronavirinae*, as virus of Middle East Respiratory Syndrome (MERS) or the Severe Acute Respiratory Syndrome (SARS) ^[1]^. In Peru, the first case of COVID-19 was identified on March 6, 2020, while the first two deaths occurred thirteen days later ^[2,3]^. Three months later, the country exceeds 260 000 cases and reports more than 8 700 deaths ^[4]^.

One of the most important characteristics of COVID-19, is the dynamics of transmission due to highly effective mechanisms. The infection agent is usually propagated through the airway or by contact of secretions; therefore, the human-to-human transmission has turned into the main dissemination path to be considered during the pandemic ^[5]^. Previous studies have determined that SARS-CoV-2 has an average reproduction number (R_0_) of 2.2, but it range from 1.4 to 6.5; however, these estimates may vary according to the study context ^[6,7]^.

Close-to-case contacts, such as family members, relatives, or friends, are those who are most at risk of contracting the infection and, therefore, can be sources of contagion for others who come in contact with them. This chain of contagion is reinforced by the fact that a percentage of the infected population can act as asymptomatic carriers of the virus, making it difficult for them to be identified by health systems ^[8]^.

In this regard, an effective way to break the chain of transmission of the SARS-CoV-2 virus is through epidemiological surveillance and follow-up of all those who were in close contact with a confirmed case ^[9,10]^. This process is called “contact tracing”. Some previous experiences have used these strategies to evaluate the transmission dynamics between COVID-19 cohabitants. A study carried out in China remarks that there was a correlation between confirmed cases in other communes in Hubei province and the number of migrants from Wuhan, who usually travel to visit their family. ^[11]^. Likewise, Liu et al. (2020), showed that family reunion events became important sources of contagion in some provinces of China, so they recommend that public health interventions should consider specific measures to reduce contact in cohabitants ^[12]^.

Additionally, previous research have found an increase between 7-10 times in secondary attack rates, when studying only the people who cohabit the home with the primary case, compared to the calculated rate when they include all contacts ^[13,14]^. However, the evidence is still divergent among regions and countries where close contact studies are taking place.

In Latin America, the weakness of the health systems and the lack of economic resources add to the difficulty of monitoring cases and contacts in this disease, this has proven to be a gravitant factor in the progression of this pandemic ^[15]^. Benitez et al, in a recent analysis in five Latin American countries, suggest that improving of a strict follow-up of contacts, as in Chile, is associated with sustained decreases in COVID-19 cases ^[16]^. Additionally, the contact tracing has been implemented late in the region, being still incomplete in countries with high mortality rates such as Brazil, Ecuador or Peru ^[17]^.

In our country, centralism is an additional factor; thus, the capital (Metropolitan Lima), concentrates approximately 60% of reported national cases. Additionally, within it, districts with high and low proportion of cases have been identified, which have varied over time ^[4]^.

Although a previous study has been found that preliminarily analyzes the dynamics of the transmission of SARS-CoV-2 in Lima ^[18]^, no analyzes have been found that evaluate the information of activities that have involved the monitoring of contact clusters narrow, as are the people who live in the same household, considering the case burden by districts of residence. Therefore, the objective of the present study is to describe the characteristics of SARS-CoV-2 infection among household members with a confirmed primary case of COVID-19 in low burden districts in Metropolitan Lima.

## MATERIALS AND METHODS

### Study design

An observational and retrospective study using epidemiological surveillance data form National Health Institute was conducted.

### Population and sample

The study population was defined as the totality of contacts with a serological test result for COVID-19. We include contacts with complete epidemiological records with IgG / IgM results, made by National Institute of Health (NIH) personnel, included in the epidemiological surveillance of households were selected. Those contacts who do not cohabit in the home of the primary case were excluded. Therefore, this study is considered a census type.

### Epidemiological surveillance

In the context of pandemic control and surveillance, the National Institute of Health (INS) carried out an epidemiological surveillance activity in households with a single primary case COVID-19 (identified by RT-PCR), between April 23 and May 2, 2020. This evaluation was carried out, on average, 13.6 ± 3.7 days after the diagnostic test.

In order to avoid inclusion of data produced by a community transmission scenario, this activity was carried out in the districts with the lowest burden of disease in each of four health areas of Metropolitan Lima, called *Directorates of Integrated Health Networks* (DIRIS, for its acronym in Spanish), until surveillance of 10 households in each DIRIS was reached. In order to that, the results of molecular tests (RT-PCR), registered until April 9^th^ on Ministry of Health’ NetLab 2.0 system, were obtained.

The identified results were classified by districts and case frequencies by DIRIS. After that, ten houses were selected, by a non-probabilistic, intentional mode, from DIRIS with the less proportion of COVID-19 cases. Two additional houses were added from DIRIS of centric zone of Lima (with more populational density). In addition, 10 households in the district of Metropolitan Lima with the highest burden at the time the surveillance began was evaluated. At the end, 52 houses were included in this study.

Subsequently, as part of the surveillance, 12 households from one of the DIRIS from centric zone of Lima were re-evaluated, on average 33.6 ± 2.7 days after the first evaluation.

The serological test used was Coretests ® COVID-19 IgM / IgG Ab Test (Core Technology Co. Ltd), a lateral flow immunochromatographic test that qualitatively detects the presence of antibodies against SARS-CoV-2, with a sensitivity and specificity reported by the manufacturer for IgM / IgG of 97.6% and 100%, respectively. Those values were verified by National Institute of Health (Peru), through laboratory evaluations, reporting a sensibility of 96.4% and a specificity of 96.0%, for IgG and IgM, respectively ^[19]^.

### Variables

The study considers a main variable called SARS-CoV-2 infection, and defined as the presence of antibodies (IgM, IgG or both) in people who had not previously being tested with a positive result (RT-PCR or serological test). The positive cases were classified in turn, according to the presence / absence of symptoms.

Likewise, information was collected on the number of members in the household, evaluation time, defined as the time in days between the issuance of the index case result and the first evaluation; and time of illness, characterized as the time in days (reported by the patient) from the onset of symptoms to the day of evaluation.

Socio-demographic characteristics of the household members (age, gender, presence of health professionals), and clinical characteristics (presence of symptoms and risk conditions) were also described. In this regard, the presence of cough, sore throat, nasal congestion, fever, general discomfort, respiratory distress, diarrhea, nausea / vomiting, headache, irritability / confusion, pain in general, among others, were considered as symptoms; while the following were included as risk conditions: age greater than or equal to 60 years, hypertension (HT), cardiovascular disease, type 2 diabetes (T2DM), obesity, asthma, chronic lung disease, chronic kidney failure (CKF), disease or immunosuppressive treatment, cancer, pregnancy or postpartum, being a health professional or others that the health personnel consider convenient to register ^[20]^.

### Statistical analysis

The descriptive statistical analysis of the data was carried out by determining the frequency, percentage, mean and standard deviation of the collected data. This evaluation was carried out in a differentiated way, expressing simple measures for the information of the subjects in general; while, to identify the values of the people within each household, average measures were used considering the variability that exists in each household according to the density of its members. Analyzes were repeated for reevaluated individuals and households to identify changes over time.

All calculations were performed with the statistical software Stata v.16.0 (Stata Corporation, College Station, Texas, USA ®), and Microsoft Excel 2016 ®.

### Ethical aspects

Due to the use of secondary data analysis for this study, in the context of an activity of epidemiological surveillance, an informed consent was no required. The confidentiality of participants personal data was preserved by using an anonymous database. Any information which permits the identification of people included was no recollected. The study has the approval of the Institutional Ethics Committee for Research from the National Institute of Health (RD N°256-2020-OGITT/INS).

## RESULTS

Records of 326 people were evaluated, in which 54.7% (n = 129) were women, with an average age of 36.2 ± 20.1. The 37.3% had some risk condition (n = 88), being the most frequent belonging to the risk age group older than 60 years (n = 35, 39.8%), followed by HT (n = 20, 22.7%) and bronchial asthma (n = 14, 15.9%). Regarding the clinical characteristics, 68.6% presented some sign and / or symptom; highlighting the presence of sore throat (49.4%), while fever and / or chills, as well as cough occurred in 41.4% of people.

Of all the subjects, 53.0% were identified as a secondary case infection, finding 15 people only with positive reaction to IgM, and 110 with reaction for both IgM and IgG. No patients were found who only exhibited a positive reaction to IgG. Among secondary cases, it was observed that 77.6% were symptomatic, the ratio of symptomatic to asymptomatic secondary cases was 3.5 (Table 1).

**Table 1.** Characteristics of the evaluated population

|  | **General individuals**<br>**(n=236)** |
| --- | --- |
| Age | 36.2 ±20.1 |
| Gender |  |
| Male | 107 (45.3%) |
| Female | 129 (54.7%) |
| District of residence based on burden of cases |  |
| Low burden | 185 (78.4%) |
| High burden | 51 (21.6%) |
| Risk condition presence |  |
| Not present | 148 (62.7%) |
| Present | 88 (37.3%) |
| Identified risk condition* |  |
| Diabetes | 13 (14.8%) |
| Chronic Arterial Hypertension | 20 (22.7%) |
| Asma | 14 (15.9%) |
| Kidney failure | 1 (1.14%) |
| Heart disease | 7 (8.0%) |
| Obesity | 9 (10.2%) |
| Pneumonia | 2 (2.3%) |
| Fibromyalgia | 1 (1.1%) |
| Anemia | 3 (3.4%) |
| Hypothyroidism | 7 (8.0%) |
| Tuberculosis | 3 (3.4%) |
| Cancer | 4 (4.6%) |
| Auto-immune disease | 1 (1.1%) |
| Health care professionals | 9 (10.2%) |
| Risk age group | 35 (39.8%) |
| Presence of sign and symptoms |  |
| Not present | 74 (31.4%) |
| Present | 162 (68.6%) |
| Time of the disease** | 11.8 ±7.5 |
| Identified sign and symptoms ** |  |
| Cough | 67 (41.4%) |
| Sore throat | 80 (49.4%) |
| Nasal congestion | 35 (21.6%) |
| Respiratory difficulty | 22 (13.6%) |
| Fever / Chill | 67 (41.4%) |
| General discomfort | 56 (34.6%) |
| Diarrhea | 27 (16.7%) |
| Nausea / vomiting | 7 (4.3%) |
| Headache | 62 (38.3%) |
| Irritability / Confusion | 4 (2.5%) |
| Pain | 1 (0.6%) |
| Anosmia | 28 (17.3%) |
| Ageusia | 31 (19.1%) |
| Test Result |  |
| Negative | 111 (47%) |
| Positive | 125 (53%) |
| Positive and Negative Ratio | 1.1 |
| Immunoglobulin Detected |  |
| IgM only | 15 (12.0%) |
| IgM / IgG | 110 (88.0%) |
| IgG only | 0 (0.0%) |
| Positive Case Type |  |
| Asymptomatic | 28 (22.4%) |
| Symptomatic | 97 (77.6%) |
| Symptomatic and Asymptomatic Positive Ratio | 3.5 |

\* The values were calculated considering only those who did present some risk condition.
\*\* The values were calculated considering only those who did present any signs and / or symptoms.

Similar age was found among people classified by results (positive / negative) and by symptoms (symptomatic / asymptomatic). The 40.2% of the symptomatic cases and 32.1% of the asymptomatic had some risk condition, being the most frequently found characteristic, belonging to the age group over 60 years, in both groups. The most frequently found signs and symptoms, in positive cases, were fever and / or chills (40.0%), sore throat (39.2%), cough (35.2%), headache (30.4%) and general malaise (28.0%). Ageusia and anosmia were present in 22.4% and 20.8% of cases, respectively (Figure 1). The type of immunoglobulin detected was similar between symptomatic and asymptomatic secondary cases (Table 2).

**Table 2.** Characteristics of people with positive and negative results, according to the presence of symptoms.

|  | Negative person |  |  | Positive person |  |  |
| --- | --- | --- | --- | --- | --- | --- |
|  | No symptoms<br>(n=46) | With symptoms<br>(n=65) | Total<br>(n=111) | No symptoms<br>(n=28) | With symptoms<br>(n=97) | Total<br>(n=125) |
| Age | 37.7 ±18.9 | 33.1 ±19.1 | 35.0 ±19.1 | 39.5 ±21.2 | 36.5 ±20.9 | 37.2 ±20.9 |
| Sex |  |  |  |  |  |  |
| Male | 22 (47.8%) | 31 (47.7%) | 53 (47.8%) | 11 (39.3%) | 43 (44.3%) | 54 (43.2%) |
| Female | 24 (52.2%) | 34 (52.3%) | 58 (52.2%) | 17 (60.7%) | 54 (55.7%) | 71 (56.8%) |
| Presence of Risk Condition |  |  |  |  |  |  |
| Present | 24 (52.2%) | 47 (77.3%) | 71 (64.0%) | 19 (67.9%) | 58 (59.8%) | 77 (61.6%) |
| Not present | 22 (47.8%) | 18 (27.7%) | 40 (36.0%) | 9 (32.1%) | 39 (40.2%) | 48 (38.4%) |
| Risk Condition |  |  |  |  |  |  |
| Anemia | 1 (2.2%) | 0 (0.0%) | 1 (0.9%) | 0 (0.0%) | 2 (2.1%) | 2 (1.6%) |
| Pneumonia history | 0 (0.0%) | 1 (1.5%) | 1 (0.9%) | 1 (3.6%) | 0 (0.0%) | 1 (0.8%) |
| Bronchial asthma | 4 (8.7%) | 2 (3.1%) | 6 (5.4%) | 1 (3.6%) | 7 (7.2%) | 8 (6.4%) |
| Cancer | 1 (2.2%) | 1 (1.5%) | 2 (1.8%) | 1 (3.6%) | 1 (1.0%) | 2 (1.6%) |
| Heart disease | 1 (2.2%) | 2 (3.1%) | 3 (2.7%) | 1 (3.6%) | 3 (3.1%) | 4 (3.2%) |
| Diabetes | 5 (10.9%) | 1 (1.5%) | 6 (5.4%) | 1 (3.6%) | 6 (6.2%) | 7 (5.6%) |
| Autoimmune disease | 0 (0.0%) | 0 (0.0%) | 0 (0.0%) | 0 (0.0%) | 1 (1.0%) | 1 (0.8%) |
| Fibromyalgia | 0 (0.0%) | 0 (0.0%) | 0 (0.0%) | 0 (0.0%) | 1 (1.0%) | 1 (0.8%) |
| Age Group at Risk | 7 (15.2%) | 5 (7.7%) | 12 (10.8%) | 6 (21.4%) | 17 (17.5%) | 23 (18.4%) |
| Chronic Arterial Hypertension | 6 (13%) | 5 (7.7%) | 11 (9.9%) | 3 (10.7%) | 6 (6.2%) | 9 (7.2%) |
| Hypothyroidism | 3 (6.5%) | 2 (3.1%) | 5 (4.5%) | 0 (0.0%) | 2 (2.1%) | 2 (1.6%) |
| Renal insufficiency | 1 (2.2%) | 0 (0.0%) | 1 (0.9%) | 0 (0.0%) | 0 (0.0%) | 0 (0.0%) |
| Obesity | 3 (6.5%) | 2 (3.1%) | 5 (4.5%) | 1 (3.6%) | 3 (3.1%) | 4 (3.2%) |
| Health professional | 1 (2.2%) | 2 (3.1%) | 3 (2.7%) | 1 (3.6%) | 5 (5.2%) | 6 (4.8%) |
| Tuberculosis | 0 (0.0%) | 2 (3.1%) | 2 (1.8%) | 0 (0.0%) | 1 (1.0%) | 1 (0.8%) |
| Immunoglobulin Detected |  |  |  |  |  |  |
| IgM only | ----- | ----- | ----- | 3 (10.7%) | 12 (12.4%) | 15 (12.0%) |
| IgM / IgG | ----- | ----- | ----- | 25 (89.3%) | 85 (87.6%) | 110 (88.0%) |
| IgG only | ----- | ----- | ----- | 0 (0.0%) | 0 (0.0%) | 0 (0.0%) |

The 236 people evaluated belonged to 52 households, finding a density of 4.5 ± 2.5 members for each household; considering the variability in the number of members per household, it was found that 54.1% of the members were women, 34.7% of the members per household had some risk condition, and 68.1% presented some sign and / or symptom. On average, 49.9% of the members of each household were identified as a secondary case for COVID-19. Of the 40 households that presented secondary cases, in 9 (22.5%) all of its members had a positive result. Additionally, on average, 39.4% of the members of each household were symptomatic secondary cases, and the ratio of finding symptomatic secondary cases was 3.8 with respect to the asymptomatic ones (Table 3).

**Table 3.** Characteristics of household composition (people per household).

| Characteristics of Household Members | Households in general (n=52) |
| --- | --- |
| Household Members | 4.5 ±2.5 |
| Age | 36.2 ±20.1 |
| Sex |  |
| Male | 45.9% |
| Female | 54.1% |
| Persons per Household with Presence of Comorbidities |  |
| Not present | 65.3% |
| Present | 34.7% |
| Persons per Household with Presence of Signs and / or Symptoms |  |
| Not present | 31.9% |
| Present | 68.1% |
| Illness Time * | 12.1 ±7.3 |
| Result of Rapid Serological Test of People per Home |  |
| Negative | 50.1% |
| Positive | 49.9% |
| People with Reactive IgM per Household |  |
| Negative | 50.1% |
| Positive | 49.9% |
| People with Reactive IgG per Household |  |
| Negative | 57.1% |
| Positive | 42.9% |
| Homes according to Positive Cases |  |
| Homes where all people were negative | 12 (23.1%) |
| Households where at least 1 person was Positive | 40 (76.9%) |
| Homes with negative and positive people | 31 (77.5%) |
| Homes where all people were positive | 9 (22.5%) |
| Homes with Positive cases according to the presence of Signs and Symptoms |  |
| Households where at least 1 person was asymptomatic positive | 11 (76.9%) |
| Households where at least 1 person was symptomatic positive | 35 (23.1%) |
| Household ratio with at least 1 symptomatic and asymptomatic positive person | 3.2 |
| Distribution of people according to their results by household |  |
| Negative Persons by Home | 50.1% |
| Asymptomatic Positive People by Home | 10.5% |
| Positive Symptomatic Persons by Home | 39.4% |
| Ratio of Symptomatic and Asymptomatic Positive People per Household | 3.8 |

The percentage values correspond to the average percentage of the characteristic of people per household.
\* The values correspond to the average of the characteristic of people per household with the presence of some sign and / or symptom.

When evaluating the characteristics of the households according to the positivity of its members, it was found that in those where all of its members were positive, 66.7% were women, while where all were negative was 55.0%. Regarding conditions of risk, a higher frequency was found as households had more positive members (Table 4).

**Table 4.** Characteristics of the composition of households (people per household) according to the results of its members.

|  | Household with all members with negative tests (n=12) | Household with negative and positive tests (n=31) | Household with all members with positive tests (n=9) |
| --- | --- | --- | --- |
| Number of people | 41 | 162 | 33 |
| Household Members | 3.4 ±2.0 | 5.2 ±2.3 | 3.7 ±3.1 |
| Age | 36.4 ±18.9 | 36.8 ±20.3 | 33.0 ±20.7 |
| Sex of the Members by Household |  |  |  |
| Male | 45.0% | 50.0% | 33.3% |
| Female | 55.0% | 50.0% | 66.7% |
| Persons per Household with Presence of Signs and / or Symptoms |  |  |  |
| Present | 52.0% | 28.6% | 16.4% |
| Not present | 48.0% | 71.4% | 83.6% |
| Illness Time * | 25.7 ±4.9 | 25.0 ±8.2 | 21.8 ±6.5 |
| Persons per Household with Risk Condition |  |  |  |
| Not present | 70.5% | 64.1% | 62.7% |
| Present | 29.5% | 35.9% | 37.3% |
| Households with a member who is a health professional | 4 | 4 | 2 |

The percentage values correspond to the average percentage of the characteristic of people per household.
\* The values correspond to the average of the characteristic of people per household with the presence of some sign and / or symptom.

The reevaluation was conducted in 40 people distributed in 12 households 33.6 ± 2.7 days after the first evaluation. The average age of the people included was 34.2 ± 17.2 years. An average of 66.8% female members per household and 39.6% people with risk condition per household were found.

On the first visit, an average of 1.9 ± 1.4 inhabitants per household had any sign and / or symptom (59.2%) of COVID-19, while in the reevaluation there were 0.9 ± 0.5 (41.6%). In the first evaluation, 1.8 ± 1.5 positive cases per household were found (57.0%), while for the re-evaluation this average was 2.0 ± 1.5 (65.6%), and the ratio of positive cases in the members per household went from 1.33 to 1.91. All IgM + IgG positive cases in the first evaluation showed positive IgM + IgG in the reevaluation.

The only case reactive only to IgM in the first evaluation, also present IgG for the second evaluation. Additionally, three new cases were identified that were initially negative and on the second visit exhibited positive reaction for IgM and IgG. The average of positive symptomatic cases per household, on the first visit, was 1.3 ± 1.4 (44.6%); and on the second visit was 0.8 ± 0.4 (37.4%). The ratio of symptomatic positives to asymptomatic ones changed from 3.60 to 1.33. (Table 5)

**Table 5.** Variation of the characteristics of people in general and people per household in the first evaluation and reevaluation.

|  | First evaluation |  | Re-evaluation |  |
| --- | --- | --- | --- | --- |
|  | Evaluation of individuals* (n=40) | Evaluation of individual per household** (n=12) | Evaluation of individuals* (n=40) | Evaluation of individual per household ** (n=12) |
| Age | 34.2 ±17.2 | ----- | ----- | ----- |
| Sex |  |  |  |  |
| Male | 14 (35.0%) | 1.2 ±0.9 (33.2%) | ----- | ----- |
| Female | 26 (65.0%) | 2.2 ±1.6 (66.8%) | ----- | ----- |
| Presence of Risk Condition |  |  |  |  |
| Present | 24 (60.0%) | 2.0 ±1.5 (60.4%) | ----- | ----- |
| Not present | 16 (40.0%) | 1.3 ±1.4 (39.6%) | ----- | ----- |
| Presence of Signs and / or Symptoms |  |  |  |  |
| Not present | 17 (42.5.0%) | 1.4 ±1.2 (40.8%) | 29 (72.5%) | 2.4 ±2.1 (58.4%) |
| Present | 23 (57.5.0%) | 1.9 ±1.4 (59.2%) | 11 (27.5%) | 0.9 ±0.5 (41.6%) |
| Test Result |  |  |  |  |
| Negative | 19 (47.5%) | 1.6 ±1.7 (43.0%) | 16 (40.0%) | 1.3 ±1.7 (34.4%) |
| Positive | 21 (52.5%) | 1.8 ±1.5 (57.0%) | 24 (60.0%) | 2.0 ±1.5 (65.6%) |
| Ratio of Positive and Negative Persons | 1.1 | 1.3 | 1.5 | 1.9 |
| Immunoglobulin Detected |  |  |  |  |
| IgM only | 1 (2.5%) | 0.1 ±0.3 (8.3%) | 0 (0.0%) | 0.0 ±0.0 (0.0%) |
| IgM / IgG | 20 (50.0%) | 1.7 ±1.6 (48.7%) | 24 (60.0%) | 2.0 ±1.5 (65.6%) |
| IgG only | 0 (0.0%) | 0.0 ±0.0 (0.0%) | 0 (0.0%) | 0.0 ±0.0 (0.0%) |
| Type of Secondary Cases |  |  |  |  |
| Asymptomatic | 5 (12.5%) | 0.4 ±0.7 (12.4%) | 14 (35.0%) | 1.2 ±1.4 (28.2%) |
| Symptomatic | 16 (40.0%) | 1.3 ±1.4 (44.6%) | 10 (25.0%) | 0.8 ±0.4 (37.4%) |
| Reason of Symptomatic and Asymptomatic Positive People | 3.2 | 3.6 | 0.7 | 1.3 |

\* Distribution of frequency and percentage of people in general.
\*\* Distribution of mean ± SD and percentage of people per household.

## DISCUSSION

This research found a secondary attack rate among cohabitants of 53.0%, which is higher than that found in other research evaluating the transmission of SARS-CoV-2 in similar clusters. The study that obtained similar results to those shown in our study was that of Wu et al. (2020), in which 148 close contacts in China were evaluated, all cohabitants of a primary case. In the publication, a secondary attack rate of 32.4% was found (95% CI 22.4% -44.4%)^[21]^.

In other countries such as China, the United States or Korea, secondary attack rates in cohabitants ranged from 4.6% to 17.0%. However, it should be considered that these estimates are affected by the sample size obtained, which was from a minimum of 151 to a maximum of 2,370 cohabitants of confirmed cases. ^[13,22–24]^. This divergence can be explained by the social and cultural differences between the countries in which these studies have been carried out, as well as the measures of social isolation and quarantine applied by the country. It should be mentioned that no history of similar studies has been found in Latin America, so the true magnitude of the influence of these factors on the progression of COVID-19 transmission in the home environment is not known.

Another explanation for these results is the time elapsed between the detection of the primary case and the secondary case, which in this investigation was an average of 13 days. Guan et al. (2020) found, in a follow-up of contacts living in the same home, that 13 days after the detection of the first case, more than half of the secondary cases had already been identified ^[25]^. Similarly, Qian et al (2020) showed 88.8% detection of secondary cases in the same household, in a follow-up carried out in China. ^[26]^.

The secondary cases had an average age of 36.1 ± 20.1 years and 54.7% were female cases. This distribution is consistent with that found by the results of the systematic review by Lovato and De Phillips (2020), which found 42.5% of male cases and a mean age of 49.1 years. ^[27]^. Likewise, it is similar to reports by the National Center for Epidemiology, Prevention and Control of Diseases (CDC-Peru), which mentions that, in Peru, 59.9% of cases are distributed in an age range between 30-59 years and 41.8% in male patients ^[28]^.

The 38.4% of positive cases reported at least one risk conditions, according to current regulations in Perú ^[29]^. The most frequent risk condition was age older or equal to 60 years (18.4%); similar to the official information of the CDC-Peru (17.3% of cases older than or equal to 60 years)^[28]^. Davies et al (2020), estimate that 69% of cases in older adults present clinical symptoms, while the susceptibility to infection decreases by half in people under 20 years of age. ^[30]^.

Other risk conditions reported in secondary cases were HT (7.2%), asthma (6.4%) and DM2 (5.6%). Previous research is contradictory regarding the frequency of cases of HT found in patients with COVID-19, being reported from 1.9% ^[31]^ to 17,4% ^[27]^; while the antecedent of asthma varied between 8.8% -12.5% ^[32,33]^, similar to that found in this research, and to that reported in Metropolitan Lima (18-19%) by other researchers ^[34,35]^. Finally, the frequency of T2DM, found in this research is consistent with that found by Tabata et al. (2020)^[31]^; although it is higher than that reported by other studies, in which the frequencies are 3%, on average ^[27,36]^.

The triad of symptoms most frequently found was fever, sore throat and cough, observed in approximately 40-50% of symptomatic positive cases. These findings are consistent with previous studies that show that fever and cough were the most frequent symptoms present in up to 80% of cases. ^[27,37]^. Bi et al.(2020) found a statistically significative relation with a PR: 3,06 [IC 95% 1,69-5,49] between the finding of fever and the detection of COVID-19 ^[22]^. Although, previous reports indicates that IgM emergence is prior of IgG, and they are positive during the first-second week after the onset of symptoms ^[38]^. Because of that, it should be considered in the current context, that the appearance of suggestive clinical characteristics, such as those previously mentioned, should lead to a rational diagnostic suspicion, with the decision to apply a test only to confirm it.

Additionally, 22.4% of the symptomatic cases presented ageusia and 20.8%, anosmia. The evidence is not yet clear regarding the frequency of these findings in cases of COVID-19, some investigations estimate its presence in more than 50% of cases ^[39]^ however, this is not supported by national evidence provided by CDC-Peru, which reports a frequency of 1.1% anosmia and 0.3% ageusia ^[28]^. The information bias that may exist must be considered, since these symptoms were not frequently consulted in the epidemiological evaluation of cases. However, it was evidenced that 92.9 and 90.3% of the contacts with these symptoms were positive to the serological test. Additionally, Patel et al. (2020) reported that 58% of cohabiting contacts in patients with anosmia and COVID-19 ^[39]^. Future studies should better assess the characteristics related to the appearance of these symptoms and transmission of SARS-CoV-2.

This research found 22.4% of positive cases asymptomatic, which is similar to the proportion of 29% reported by CDC-Peru ^[28]^. Likewise, this is consistent with what was found in other contact surveillance studies, such as the one carried out by Bi et al (2020), in China, where 20% of secondary cases were asymptomatic ^[22]^; and by Hao-Yuan et al (2020), in Taiwan, who calculated 18.2% of secondary asymptomatic cases ^[40]^.

On the re-evaluation of cases, it was found that although the frequency of people with symptoms fell, the ratio of positive cases rose from 1.33 to 1.91. This is consistent with the reevaluation time (more than 30 days on average), since the sensitivity of the detection of antibodies in the population increases proportionally to the time of illness ^[41]^. Despite this, no significant number of seroconversions were found in people who were negative in the first evaluation, and the IgM persisted positive.

It should be noted that in this research, households have also been characterized as units of measurement, establishing corrections in epidemiological indicators for household density. This consideration is extremely important if a contact study is carried out in specific conglomerates such as houses, especially in a non-random selection mode, as in this case. Thus, in 23.1% of the homes evaluated, no positive case was found, with an average density of 4.5 ± 2.5 people per home and 3.7 ± 3.1 people in the homes where all the members were positive. This density was similar and would not explain the absence or increased infection of contacts in these households. Jing et al (2020), carried out a similar experience, finding 65% of households without positive cases, with a median of members per household of 6 (4-10) people; however, they did not analyze the characteristics of households with positive cases ^[24]^.

Additionally, and as expected, there was found an important difference between the percentage of people with symptoms in households with all positive contacts, compared to households with all negative contacts (83.6 vs. 48%). Regarding the differences between households in districts with low case burden and households in districts with high burden, was evidenced as expected in the first group the percentage of contacts with signs and symptoms, and of contacts with positive results is lower (65.5 vs. 79.2% and 48.5 vs. 55.6%, respectively).

This research has important limitations to consider: the selection of the households in the epidemiological surveillance activity was carried out for convenience, so the results presented cannot be extrapolated to the general population. Likewise, there was no component of temporality for all the cases, which does not allow establishing whether the cases called “asymptomatic” were actually pre-symptomatic cases. The reevaluation activity could not be carried out in all the households initially included, which adds an important selection bias and reduces the external validity of the conclusions that can be obtained from these data. Despite this, due to not finding similar antecedents in Latin America, this study presents results that can serve as a basis for future research that generates knowledge about the dynamics of household transmission of SARS-CoV-2.

Although the serological tests used in the study have the endorsement of the state regulatory authority (INS) through laboratory evaluations; in Peru, there is only one field study that uses this type of test for this purpose; however, this research uses another brand (Zhejang Orient Gene ®), and does not define diagnostic performance values.

Finally, it is concluded that having a primary case of COVID-19 in the home, the secondary attack rate of this infection is 53%; however, in 23% of the homes evaluated there was no positive cases, beyond the primary case. The epidemiological and clinical characteristics found in this study were in accordance with what has already been reported in other international series. Similarly, the proportion of asymptomatic patients found (22.4%) is consistent with that evidenced by previous publications and the national epidemiological data. Likewise, the persistence of positive IgM is evident in the reevaluation of cases on average 30 days later.

## Data Availability

The database is not freely available for lectors, but any contact could be done sending an email to corresponsal author.

## Author’s contribution

YAB, GS, JA and CC have participated in the conception and design of the manuscript. FC and AJ participated in the data collection, GS and YAB participated in the statistical analysis. All the authors participated in the interpretation of the data, production and review of the manuscript, final approval and are responsible for the final content.

## Acknowledgment

To Joel Roque Hernandez, Lenin Rueda Torres and Lilyana Collazos for their technical collaboration during data recollection. To Duilio Fuentes Delgado for his recommendations and technical assessment during the process of approval of the research protocol.

## Notes

### Competing Interest Statement

The authors have declared no competing interest.

### Funding Statement

The epidemiological activity was funded by National Institute of Health (Peru). This secondary data base study was self-funded.

### Author Declarations

The study has the approval of the Institutional Ethics Committee for Research of the National Institute of Health (Peru).

### Summary of Updates

Section of Methods updated to clarify; two more references were added in introduction, and Figure 1 were supressed after first peer-review round.

